# Complement expression in human atherosclerotic plaques and blood cells in patients with coronary artery disease

**DOI:** 10.1101/2025.08.08.25333332

**Authors:** Eero Teppo, A. Inkeri Lokki, Emma Raitoharju, Harlan Barker, Pashupati P. Mishra, Saara Marttila, Jaakko Laaksonen, Leo-Pekka Lyytikäinen, Nina Mononen, Ivana Kholová, Seppo Parkkila, Ari Mennander, Mika Kähönen, Seppo Meri, Niku Oksala, Terho Lehtimäki

## Abstract

**Background:** Inflammatory processes are a key cause of atherosclerosis and cardiovascular diseases. Complement system has been implicated but evidence is less clear. We tested whether atherosclerosis severity, plaque types, and vascular region are associated with mRNA expression of genes encoding proteins of the complement system and investigated the cell-specific expression of the most differentially expressed transcripts in plaque macrophages, fibroblasts, and endothelial, smooth muscle, Schwann, mast, plasma, T-, and B-cells.

**Methods:** Total mRNA was isolated, and gene expression analyzed from 29 carotid, 15 abdominal aortic, and 24 femoral plaque samples, and 28 atherosclerosis-free control left internal thoracic artery samples of 95 patients, as well as from 97 whole blood and 97 peripheral mononuclear cell samples of 97 patients. Genome-wide transcriptomic analyses were done using RNA bead microarray platforms. Differential expression was compared between plaques with normal arteries, unstable with stable plaques, as well as CAD patients with CAD-free patients.

**Results:** A total of 33 out of 90 (37%) transcripts of the complement system were differentially expressed in atherosclerotic plaques as compared to histologically normal arteries. In aortic, carotid, and femoral plaques 38, 36 and 29 transcripts, respectively, were differentially expressed, of which 25 were shared by the plaques of all arterial beds. Among the most interesting gene-level findings, we observed that transcripts of the integrin gene ITGB2 are highly upregulated mostly in macrophages and T cells while having top centrality in the network and top enriched genesets related to cell junctions.

**Conclusions:** This is the first study exploring the association of local complement expression across multiple arterial beds and histologically diverse plaque samples. The local effect of complement-mediated inflammation is indicated in inflammatory plaque tissue but not in smooth muscle cell-dominated plaques. Taken together, gene expression of the complement system components in artery cells is associated with advanced atherosclerosis.

## Introduction

Cardiovascular disease (CVD) is the leading cause of mortality worldwide, with most CVD-related deaths resulting from myocardial infarction or stroke^1^. The main underlying cause of thrombosis and cardiovascular events is atherosclerosis, a complex inflammatory disease in which extracellular lipids and fibrotic matrix accumulate in plaques^2^.

The complement system consists of 1) pattern recognition molecules, such as C1q, MBL, properdin, P-selectin, and ficolins; 2) an enzyme cascade, including C3, FB, FD, C3 convertase, C4, C5, C5 convertase, and C7; 3) an amplification loop driven by the balance of C3b interaction and breakdown; 4) anaphylatoxins C3a, C5a, and sTCC; 5) anaphylotoxin receptors; and 6) the complement regulators DAF, MCP, C4bp, factor H, CD59, CR3, clusterin, and vitronectin. The alternative pathway has always low activity as a surveillance measure, but the classical (C1q) and lectin (MBL) pathways are typically activated in response to immune complexes or pathogens, respectively. All pathways of complement activation ultimately lead to the formation of a porous cell destroying structure. 7) Activation of the membrane attack complex (MAC) by terminal pathway of the complement system causes opsonization and destruction of pathogens; recruitment of immune cells; phagocytosis; and activation of adaptive immunity, leukocytes, endothelial cells, and platelets, as well as elimination of dying cells, antigens, and immune complexes. Consequently, a dysfunctional or dysregulated complement system is associated with the risk of infection, autoimmunity, chronic inflammation, thrombotic microangiopathy, graft rejection, and cancer^3–6^.

Many components of the complement system, including deposits of C5b-9 complexes in coronary artery plaques, have been previously detected in atherosclerotic lesions^7^. Furthermore, at least the overexpression of C1r, C1s, C4, and C7 mRNAs have been observed in aortic plaques, as has the expression of C1q in arterial wall dendritic cells, macrophages, foam cells, and neovascular endothelial cells in various arterial beds^8,9^. A high complement concentration in plasma seems to be associated with a high concentration within the arterial wall. In early lesions of the femoral and iliac arteries, vitronectin colocalizes with thickening and fibrosis, and similarly, in coronary arteries, the absence of factor H is associated with C5b-9 deposits^7,10^. At the proteomic level, the quantity of proteins related to hemostasis, lipid metabolism, and the complement system are positively associated with early atherosclerosis progression in human abdominal aortas. Interestingly, C5, DAF, and CD59 (MAC) mRNA levels were unchanged, suggesting accumulation from extravascular sources. Furthermore, C5 plasma levels provided prognostic value when basic regression models were used to predict atherosclerotic imaging features in two cohorts^11^. While much of the complement protein is likely derived from the liver, intravascular synthesis still likely has an important fine-tuning effect. Although less studied, local complement expression has been observed in other diseases as well as plaque cells, and a few paracrine, autocrine, and intracellular mechanisms have been investigated in detail^12^.

In the present study, we aimed to further describe the function of the complement system in advanced human atherosclerotic lesions in major vascular beds by studying the expression of nuclear-encoded mRNA transcripts in vascular samples from different anatomic locations and in blood samples from patients with cardiovascular disease, all of which were collected in the ongoing Tampere Vascular Study (TVS). We hypothesized that the mRNA expression profiles of complement system proteins are associated with the extent of atherosclerosis in arteries and patients and that there are differences in the expression profiles between vascular beds.

## Methods

### Vascular sample collection

Vascular sample series from the TVS^13,14^, including the femoral arteries, carotid arteries, and abdominal aortas, were obtained during open vascular procedures in the Division of Vascular Surgery and Heart Center of the Tampere University Hospital from 2005--2009 from patients fulfilling the following inclusion criteria: (1) carotid endarterectomy because of asymptomatic or symptomatic and hemodynamically significant (over 70%) carotid stenosis or (2) femoral or (3) aortic endarterectomy with aortoiliac or aortobifemoral bypass because of symptomatic peripheral arterial disease. Left internal thoracic artery (LITA) samples serving as controls were obtained during coronary artery bypass surgery because of symptomatic CAD. An exclusion criterion was a patient’s refusal to participate in the study.

### Whole blood and circulating monocyte fraction collection

TVS whole blood and monocyte collections were performed in 2008 from an angiographically verified sample of patients, which was selected from a larger population-based cross-sectional study^15^ comprising 4,131 patients subjected to an exercise test between October 2001 and December 2008 at the Tampere University Hospital and thereafter treated according to the Finnish Current Care Guidelines.

### Histological classification of atherosclerotic plaques

All vascular tissue samples were fixed in 10% formalin, processed and embedded in paraffin. Four- µm sections were stained with hematoxylin-eosin and classified microscopically by a senior cardiovascular pathologist (I.K.) according to the American Heart Association (AHA) atherosclerosis plaque definition and lesion type classification scheme^16^. Type V and VI atherosclerotic lesions were further histologically classified as stable or unstable according to the presence of fissures, rupture, hemorrhage, or thrombosis. All the LITA samples were histologically normal.

### Measurement of arterial, whole blood, and mononuclear cell mRNA expression

RNA isolation from vascular and blood samples was performed following the manufacturer’s instructions as follows. Fresh vascular tissue samples were soaked in an RNA stabilization solution (RNALater by Ambion Inc., Austin, TX, USA) and homogenized (Ultra-Turrax® T80 homogenizer by IKA, Staufen, Germany). Later, vascular RNA was isolated from the samples with an RNA isolation kit (TRIzol by Invitrogen, Carlsbad, CA, USA; the miRNeasy Mini-Kit by Qiagen, Hilden, Germany; the RNeasy Kit with DNAse Set by Qiagen, Valencia, CA, USA; and the RNase-Free DNase Set by Qiagen, Valencia, CA, USA). Similarly, whole-blood RNA samples were produced from blood samples via an RNA isolation kit (PAXgene tubes by BD, Franklin Lakes, NJ, and PAXgene Blood RNA Kit with DNAse Set by Qiagen). Mononuclear cells were isolated from whole blood samples via epichlorohydrin-copolymerized sucrose and diatrizoate sodium-based density gradient centrifugation (Ficoll-Paque by Amersham Pharmacia Biotech UK Limited, Buckinghamshire, England), and RNA was isolated from the mononuclear cell fraction via an RNA isolation kit (RNeasy Mini Kit by Qiagen). The resulting RNA concentrations were checked via a spectrophotometer (BioPhotometer by Eppendorf, Wesseling-Berzdorf, Germany), and vascular RNA quality was assessed via electrophoresis (Agilent RNA 6000 Nano Kit by Agilent, Santa Clara, CA, USA). Finally, all the RNA samples were stored at -80°C.

RNA expression was quantified from the stored RNA samples following the manufacturer’s instructions as follows. First, RNA concentrations were checked with a spectrophotometer (Nanodrop ND-1000 by Nanodrop Technologies, Wilmington, DE, USA). Next, from each RNA sample, 200 ng of RNA was reverse transcribed to cDNA and transcribed back to cRNA with an RNA amplification kit (Illumina RNA Amplification Kit by Ambion, Inc., Austin, TX, USA), and the RNA was labeled with biotin-11-deoxy-UTP (PerkinElmer Life And Analytical Sciences, Inc., Boston, MA, USA) for 14 hours in vitro. The resulting cRNA concentration was checked again via a spectrophotometer, and cRNA quality was checked via electrophoresis (Expert Automated Electrophoresis System and RNA StdSens Analysis Kit by Bio-Rad Laboratories, Inc., Hercules, CA, USA). For each vascular RNA sample, 1.500 ng of cRNA was hybridized for 18 hours at +55°C onto a microarray (for mononuclear cell RNA, the Sentrix Human-8 Expression BeadChip from Illumina, San Diego, CA, USA; for vascular and whole blood RNA, the HumanHT-12 v3 Expression BeadChip from Illumina, San Diego, CA, USA) and labeled with 1 µg/ml cyanine-3- streptavidin (Amersham Biosciences, Piscataway, NJ, USA). Finally, the label intensities for each probe were measured for each array (sample) via a laser-detection scanner and image processing software (Illumina iScan and GenomeStudio by Illumina, San Diego, CA, USA).

### Measurement of patient characteristics

Patient history data are based on hospital records and patient interviews. Age, sex, weight, height, smoking history (never/ex-smoker/current smoker), history of statin use, and history of diagnoses, including type 2 diabetes, type 1 diabetes, CVD risk factors, hypercholesterolemia, hypertension, and myocardial infarction, were measured directly after participation via a questionnaire by healthcare personnel. Data on whole blood and mononuclear samples from patients were collected between October 2001 and the end of December 2004. CAD was defined primarily as having over 50% stenosis in coronary angiography, and the angiography results themselves were extracted from the participants’ electronic health records. Finally, a few missing angiography-based CAD values were imputed by using a symptom-based prediction of coronary heart disease, or if both were missing, patients with a history of myocardial infarction were assumed to have CAD.

### Microarray data analysis

All microarray data-related analyses were performed with R version 4.2 ^17^.

### Microarray preprocessing

For each sample, the background intensity was subtracted, and the data were exported from the Illumina GenomeStudio software, imported into R, log_2_-transformed, and normalized via a method based on locally estimated scatterplot smoothing (LOESS) implemented in the lumi R package^18^. LOESS normalization was used because, in a previous study, LOESS-based microarray differential expression results had the best agreement with qRT‒PCR-based results for the same arterial samples and genes^19^. Next, data quality was controlled, including the detection of outlier arrays on the basis of a low number of robustly expressed genes and hierarchical clustering. A total of 96 RNA samples (67 plaque samples, 28 LITA samples) fulfilled the quality criteria, whereas 6 RNA samples were discarded. Next, from all the arteries, only the microarray data for the abdominal aorta, femoral artery, and carotid artery samples were included in the present study. For RNA transcripts with multiple probes, the probe with the largest median intensity over the included samples was selected to represent the transcript’s expression. All of the studied RNA transcripts (1–3 transcripts per gene) are listed in Supplementary Table S1. Genes were selected for the present analysis on the basis of Gene Ontology^20^ annotation of cellular components (Supplementary Table S2).

### Patient and histology analysis

The relevant clinical characteristics of the patients, i.e., age, sex, BMI, CAD, history of myocardial infarction, cerebrovascular disease, peripheral vascular disease, diabetes type I and II, hypercholesterolemia, hypertension, statin use, and smoking, were summarized (percentages and medians with ranges), and differences between cases (AHA I-VI vascular sample or CAD) and controls (normal artery sample or CAD-free) were tested via the Mann‒Whitney U test for continuous variables (age and body mass index) and Fisher’s exact test for binary variables with at least one observation in each cell. A few missing observations were removed within each test. The histological characteristics of the vascular samples, i.e., AHA classification (normal or I-VI, including subclasses Va-Vc) and the presence or lack of fissures, thrombosis, or hemorrhage, were also summarized within each type of artery (carotid, femoral, abdominal aorta, or LITA).

### Differential expression analysis

Differential expression analysis was performed via the Mann‒ Whitney U test^21^ with Benjamini‒Hochberg p-value adjustment^22^ over the RNA transcripts. This method is robust since it makes few assumptions about the data. To facilitate interpretation, point estimates were back-transformed, i.e., differences in pseudomedians of log_2_ values were transformed into fold-change values, and downregulation was presented as negative numbers. Transcripts with fold changes greater than 1.5/-1.5 and adjusted p-values less than 0.05 were interpreted as differentially expressed. For each RNA transcript, seven comparisons were made: 1) RNA expression in histologically normal vs. AHA type I-VI atherosclerotic plaque samples; 2) type AHA I-IV vs. V-VI; 3-5) stable vs. unstable, thrombotic, or hemorrhaged plaque samples; and 6-7) CAD vs. CAD-free patient whole blood and mononuclear cell samples. In addition, the primary comparison (AHA I-VI plaque vs. normal artery) was replicated using subgroups of vascular beds, i.e., carotid, femoral, or abdominal aortae, compared with the LITA. The results from different vascular beds were compared in two ways: 1) the number of differentially expressed transcripts in each vascular bed was compared in a Venn diagram, and 2) simple linear regression was used to predict the expression of all differentially expressed (AHA I-VI vs. normal) transcripts in plaques on the basis of the vascular bed, and the resulting coefficients were visualized in a volcano plot. Next, at most, the ten most upregulated and ten most downregulated genes were selected by ranking these transcripts by their adjusted p-values (AHA I-VI plaque vs. normal artery). In addition, the role of these top atherosclerosis-associated genes as predictors of angiographically verified CAD was investigated in whole blood or mononuclear cells via logistic regression analysis with k nearest neighbor imputation and Benjamini-Hochberg p-value adjustment over the transcripts.

### Marker correlation analysis

Pearson correlations and t-tests (WGCNA::corAndPvalue) with Benjamini-Hochberg p-value adjustment were used on the plaque (AHA I--VI) samples to measure the associations of the studied transcripts with the transcripts of 55 marker genes (Supplementary Table S2), which were selected based on previous work^23^. More specifically, correlations were analyzed with three selected markers of inflammation, three markers of smooth muscle cells, 25 markers of SMC-rich plaques, and 10 and 14 markers of M1 and M2 macrophages, respectively.

### Cluster analysis

To identify global patterns or modules of expression, two analyses were performed: Uniform Manifold Approximation and Projection (UMAP) on the artery, whole blood, and mononuclear samples and Topological Overlap Matrix (TOM) -based Weighted Graph Correlation Network Analysis (WGCNA) on the transcripts in the artery samples. The UMAP analysis was computed via 10 nearest neighbors, the TOM was based on exponentiated Pearson correlations so that a few powers were used, and the final network was chosen visually based on evaluation metrics presented in Supplementary Figure S1.

### Gene set analysis

GO term, KEGG pathway, and Mitocarta 3.0 pathway enrichment were analyzed with modified Gene Set Z-scoring (mGSZ) with Benjamini-Hochberg p-value adjustment and Fisher’s exact tests. In all analyses, all 90 transcripts formed the reference group, and only genesets with at least five transcripts were included. As enrichment is measured only within the studied system, the interpretation is slightly different from that of the usual genome-wide analysis. In Fisher’s exact tests, at least one observation was required in each cell, and transcripts with a fold change greater than -1.5/1.5 and an FDR<0.05 in the primary comparison of AHA I-VI plaques with normal arteries were defined as differentially expressed. Gene sets and pathways with FDRs<0.25, as suggested by ^24^, were interpreted to be significantly enriched and differentially expressed. Finally, these analyses were replicated using subgroups of arteries, i.e., carotid, femoral, or abdominal aorta vs. LITA, and compared for evidence of vascular bed heterogeneity.

### Analysis of the cell-specific expression of the top genes via single-cell RNA sequencing data

All single-cell data analyses were performed with Python version 3.7 and the Scanpy library^25^. The Python script can be found in the supplemental files.

### Plaque RNA-sequencing data

The transcriptomics data for our cell-specific analyses were derived from the public open data source Gene Express Omnibus (GEO)^26^. An experiment by^27^ performed scRNA-Seq analysis of human atherosclerotic lesions with publicly available data (GSE131778). These data contain read counts for 11756 human cells derived from 3–6 coronary artery plaque sections of four patients and 20431 genes and were downloaded for the present secondary analysis.

### Preprocessing

The cells were filtered by read count (1600–17000), gene read count fraction (less than 20%), and total number of genes expressed (over 300). Genes were filtered according to the number of cells expressing them (over 10). Cell and gene expression filtering resulted in a total of 10,884 cells and 17,990 genes carried forward for subsequent analyses. Counts were normalized to a million per cell (CPM) and natural log-transformed. The top 4,000 highly variable genes were extracted via the method of Cell Ranger^28^. Finally, the dimensionality was reduced to 50 via principal component analysis (PCA).

### Cell clustering and annotation

UMAP was used to compute an adjacency matrix (graph), and then Louvain clustering (resolution 0.25) was computed to identify subgroups within the scRNA-Seq data. Automated cell type annotation was performed with the SCSA tool (A Cell Type Annotation Tool for Single-Cell RNA-seq Data)^29^, which uses a modified human cell type marker gene library comprising data from the default CellMarker database^30^ as well as from the PanglaoDB database^31^. The predicted macrophage subgroup was separated for independent subclustering analysis via the same steps except that only 1,000 highly variable genes were included. Finally, force-directed graph representations were drawn with ForceAtlas2 for all cells and macrophages.

### Target gene expression in different cell types

For all cell types, the mean expression and fraction of expressing cells were calculated for the top DEGs, and the results were visualized in a dot plot.

## Results

### Patient and sample characteristics

We analyzed the data of 95 participants who provided 96 vascular samples, including 68 plaque samples from 15 abdominal aortas, 29 carotid arteries, and 24 femoral arteries, as well as 28 samples from LITA arteries. We also analyzed data from 97 participants who provided 97 mononuclear cell and 97 whole blood samples, including 52 whole blood and mononuclear cell RNA samples from CAD patients and 45 from patients without CAD.

The characteristics of the participants are summarized in Table 1. The participants had a median age of 69 years (range 40–91) and 72% male in the vascular study group and a median age of 57 years (range 38–76) and 63% male in the monocyte/whole blood study group at the time of recruitment. On the basis of the hypothesis tests, the source population of the artery case patients probably had many more PADs, fewer myocardial infarctions and CAD, and slightly lower BMIs than did the source population of the control patients; similarly, the blood case patients probably used many more statins and had more myocardial infarctions and hypercholesterolemia. These differences are expected from the study design, and the sensitivity of the differential expression results to these variables (in addition to age, sex, and BMI) was studied.

**Table 1.** Demographics and risk factors for the study patients. The summary measures for age and BMI are the median and range. Categorical data were compared with Fisher’s exact test, and continuous data were compared with the Mann‒Whitney U test. Abbreviations: LITA, left internal thoracic artery; BMI, body mass index; HC, hypercholesterolemia; CAD, coronary artery disease; MI, myocardial infarction; CeVD, cerebrovascular disease; PVD, peripheral vascular disease.

| Variable | Patients with plaques | Patients with normal LITA | Artery P value | CAD patients | CAD free patients | Blood P value |
| --- | --- | --- | --- | --- | --- | --- |
|  | n = 67 | n = 28 |  | n = 52 | n = 45 |  |
| Age (Years) | 70 (45–91) | 69 (40–81) | 0.26 | 56 (39–75) | 57 (38–76) | 0.84 |
| BMI (kg/(m<sup>2</sup>)) | 26 (17–35) | 28 (23–47) | 0.043 | 27 (19–38) | 27 (19–37) | 0.69 |
| CAD | 67% | 100% | - | 100% | 0% | - |
| CeVD | 52% | 14% | 5.7e-04 | 12% | 13% | 1 |
| DM1 | 0% | 3.4% | - | 1.9% | 2.2% | 1 |
| DM2 | 18% | 24% | 0.58 | 15% | 6.7% | 0.21 |
| HBP | 85% | 97% | 0.16 | 96% | 87% | 0.14 |
| HC | 69% | 86% | 0.12 | 77% | 51% | 0.011 |
| Male | 67% | 83% | 0.14 | 62% | 64% | 0.83 |
| MI | 15% | 39% | 0.014 | 58% | 24% | 0.0011 |
| PAD | 55% | 7.1% | 8.8e-06 | 2% | 2.2% | 1 |
| Smoked before | 45% | 45% | 1 | 27% | 27% | 1 |
| Smoked never | 22% | 34% | 0.21 | 35% | 47% | 0.3 |
| Smoking now | 33% | 21% | 0.32 | 38% | 27% | 0.28 |
| Statin | 73% | 83% | 0.44 | 75% | 29% | 1.2e-05 |

The histological characteristics of the artery samples are summarized in Supplementary Table S4. Most of the plaque samples were complicated Type VI (44%) or fibrous Type V (29%), whereas 5.9% were atheroma Type IV and 4.4% were intermediate Type III. In terms of stability, approximately 43% of the measured plaque samples were stable, another 26% were hemorrhaged only, 12% had hemorrhage thrombosis, 10% had thrombosis only, one sample (1,7%) had fissures only, and one (1,7%) hemorrhage-fissure. None of the histologically analyzed LITA samples were abnormal, as expected, and all except 1 of the non-LITA samples were histologically abnormal. This explains the apparent discrepancy in the number of LITA samples and patients with normal vascular samples.

### Differential expression analysis between non-atherosclerotic normal vessels and atherosclerotic plaques

Differential expression analyses between histologically normal vessels and all atherosclerotic plaques (AHA types I-VI) are summarized in Table 2 and Figure 1. Comparisons of advanced plaques with stable plaques for the top genes are presented in Supplementary Figure S2 and for all genes in Figure S3. The regression results for the top transcripts are shown in Supplementary Table S5.

**Figure 1.**
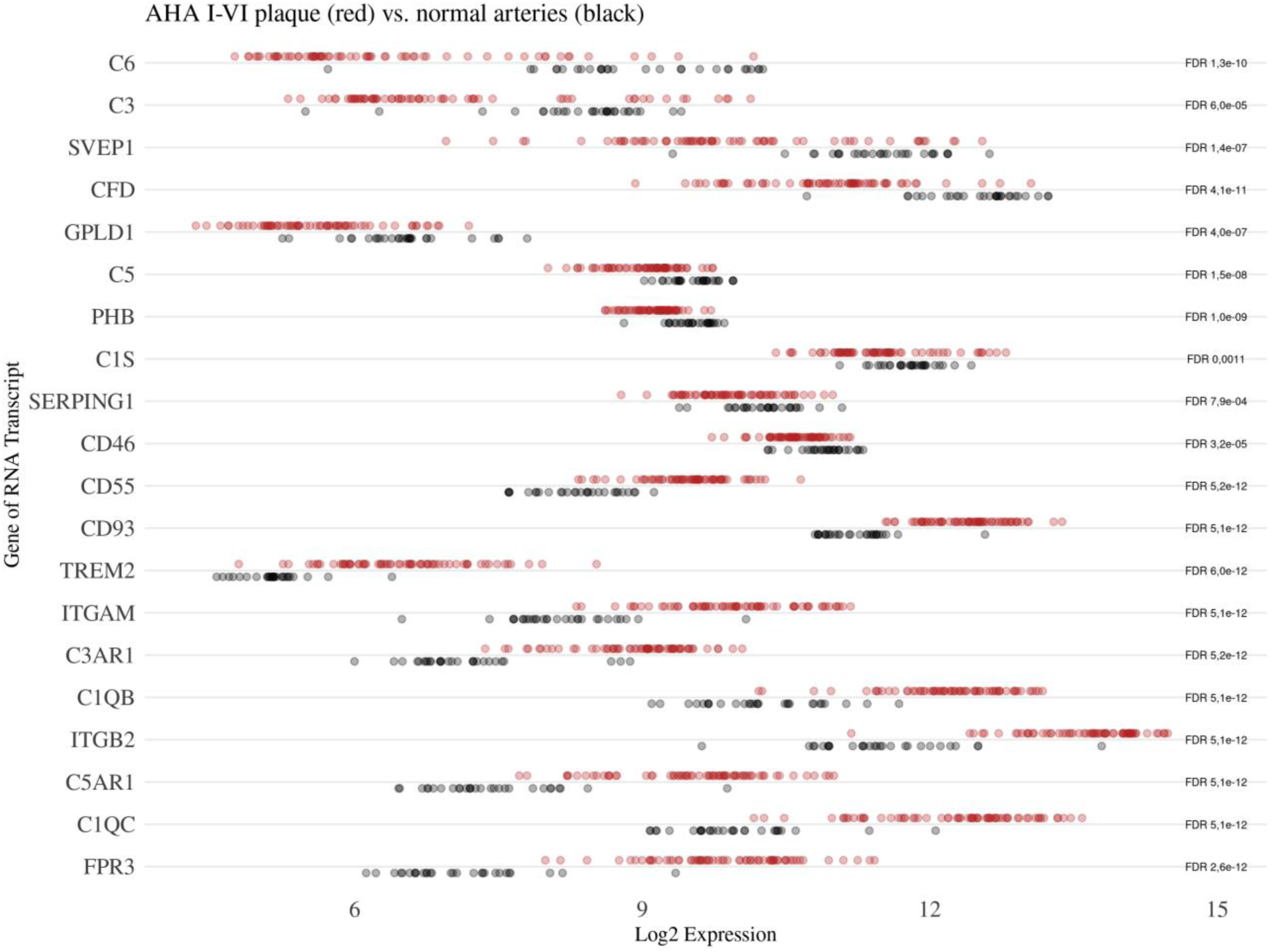
Top transcripts showing differential expression between histologically normal and atherosclerotic plaques with lesion types I-V.

**Table 2.**
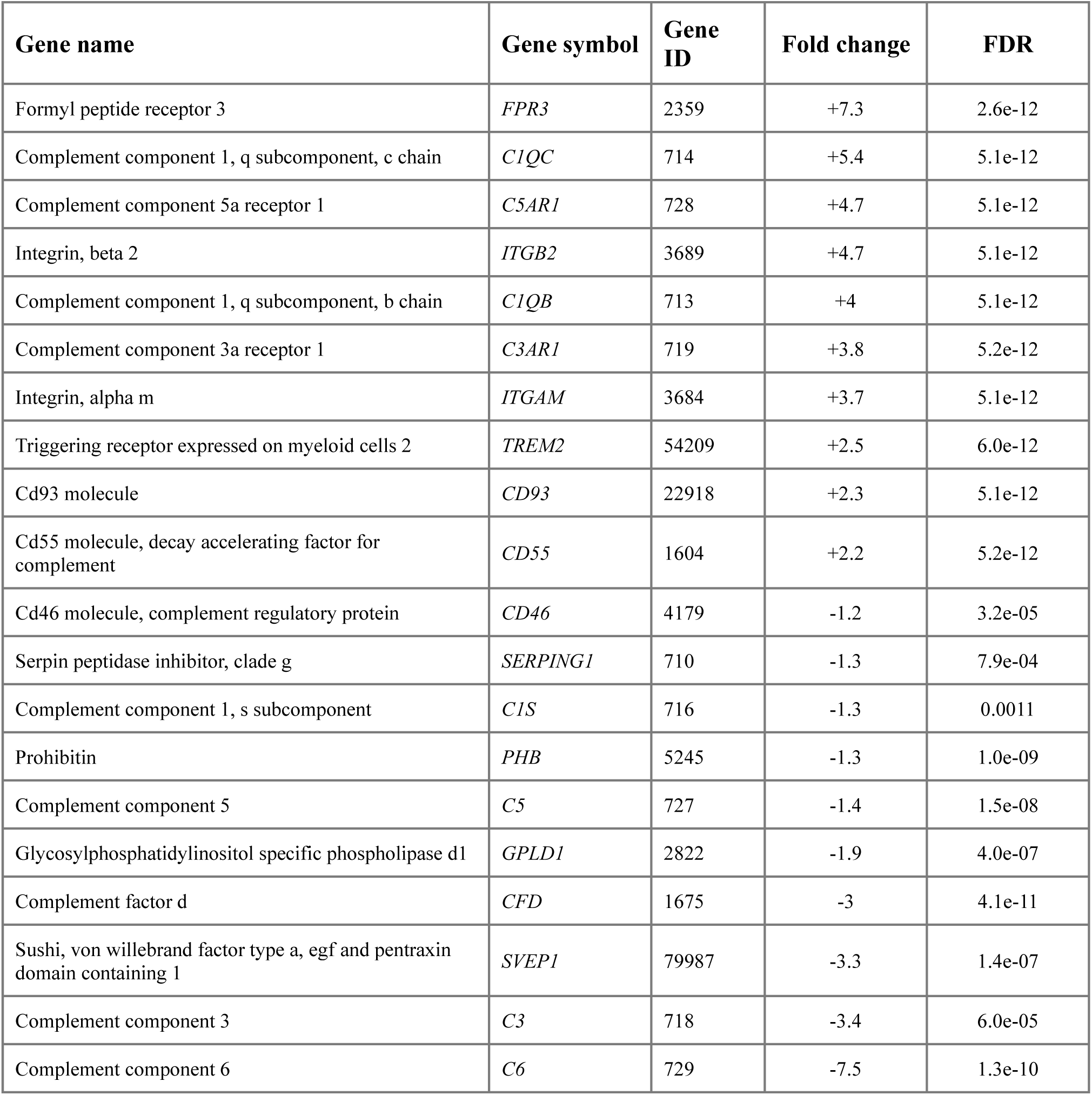
The ten most upregulated and downregulated transcripts according to the false discovery rate when comparing non-atherosclerotic normal vessels and atherosclerotic plaques (AHA I-VI: American Heart Association Stary classification scheme of atherosclerotic lesions; *Type II*, Fatty streaks; *Type III*, Intermediate lesion; *Type IV*, Atheroma; *Type Va*, Fibroatheroma; *Type Vb*, Calcific lesion; *Type Vc*, Fibrotic lesion; *Type VI*, Complicated lesion; *Type VIa*, Surface disruption/Fissure; *Type VIb*, Hematoma/Hemorrhage; *Type VIc*, Thrombosis). At most, 10 up- and 10 downregulated genes were selected, minimizing FDR values. FDR, false discovery rate (Benjamini‒Hochberg).

A total of 33 out of 90 (37%) transcripts were significantly differentially expressed between atherosclerotic plaques (AHA types I-VI) and histologically normal samples (Supplementary Table S1). Complement transcripts are mostly upregulated, but a few genes, such as *C6*, *C3*, and *CFD,* are also highly downregulated.

In addition, only a few transcripts were differentially expressed when comparing stable and thrombotic plaques (*CFP*, *CFD*, *C7*, RGCC/C13orf15), and none were statistically significant when comparing AHA type V-VI plaques with AHA type I-IV plaques or stable plaques with unstable, fissured, or hemorrhaged plaques. After adjusting for clinical features, four transcripts (*C1S*, *C3*, *FPR3*, and *SERPING1*) were statistically non-significant at the 5% level in the “extra” model (Supplementary Table S5), although all p-values were strongly increased by modeling.

### Gene expression differences between healthy control vessels and plaques derived from different vascular regions

Differential expression analyses between histologically non-atherosclerotic controls and carotid, aorta, and femoral artery plaques are summarized in Figure 2 and Supplementary Table S6.

**Figure 2.**
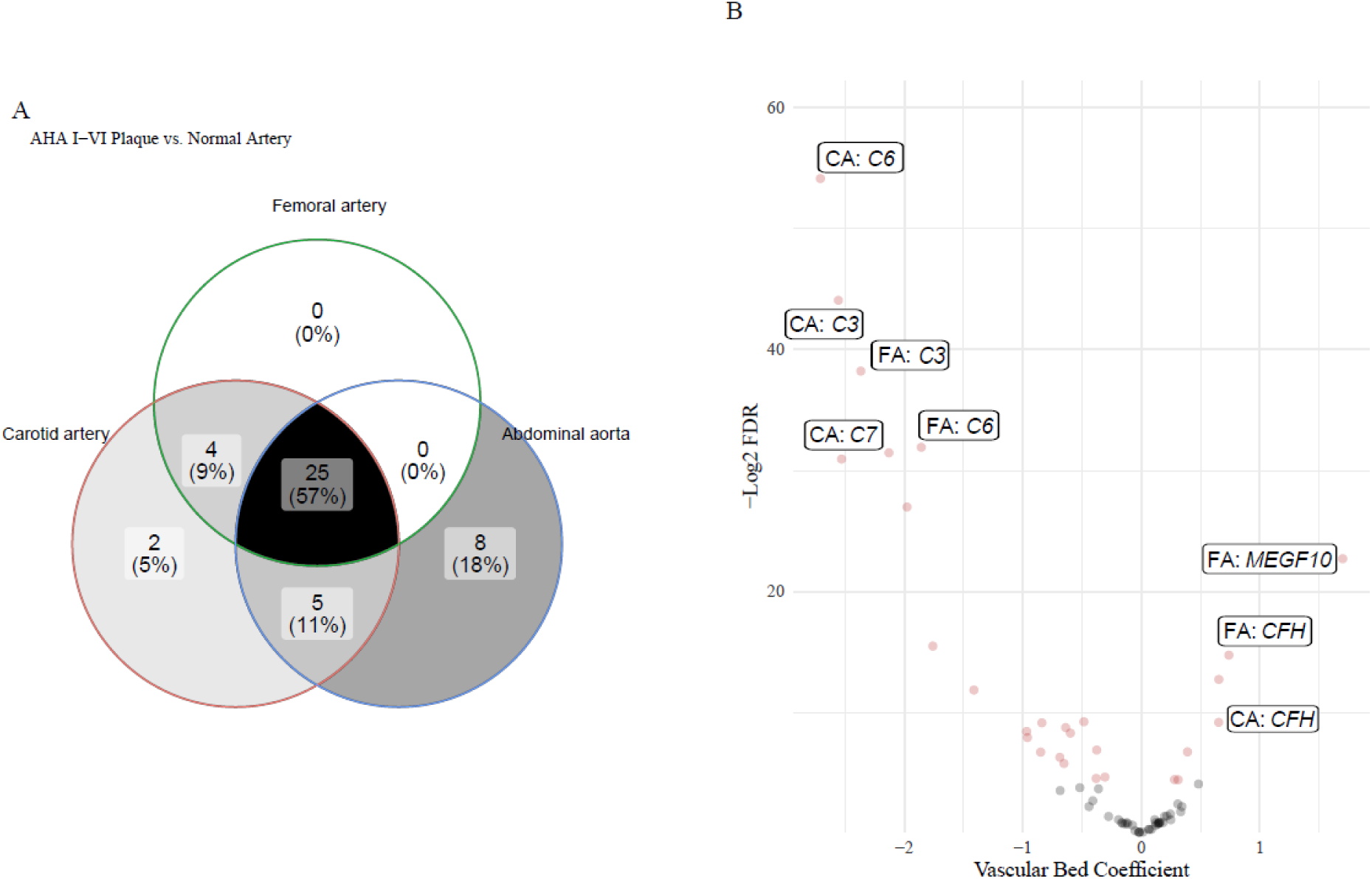
A. Venn diagram showing the number of differentially expressed transcripts in each subset, comparing AHA I-VI plaques from carotid arteries, femoral arteries, or abdominal aortas with normal arteries from LITAs. B. A volcano plot presenting all point estimates and p values of coefficients of linear regression models that predict the expression of each differentially expressed transcript on the basis of the vascular bed of the artery samples (the abdominal aorta is the reference). The color represents the multiplicity-adjusted (Benjamini‒Hochberg) statistical significance of the coefficient at the 0.05 level. A few of the most heterogeneous transcripts are labeled with the symbol of their gene. CA, carotid artery; FA, femoral artery

We investigated vascular regional differences and found minor differences in gene expression between plaques taken from different types of arteries from different vascular regions. In abdominal, carotid and femoral plaques, 38, 36 and 29 genes, respectively, were differentially expressed, of which 25 were shared by all of the arterial beds. The expression of 2 (2,2%; *SUSD4* and *PTX3*) genes in carotid plaques, 8 (8,9%) genes in aortic plaques, and 0 (0%) genes in femoral plaques were unique to our analysis. A few genes, including *C6*, *C3*, and *C7*, are expressed very differently depending on the artery. For example, *C6* has a very high expression difference in the carotid arteries at approximately -12 (p = 10E-9) compared with the abdominal aorta at -1,8 (p = 10E-2).

### Differential expression analysis between angiographically verified CAD patients and controls

In neither whole blood nor blood mononuclear cell samples were any of the transcripts significantly differentially expressed by coronary artery disease status (yes/no) after multiple testing adjustment of p-values. Similarly, after two sets of clinical features were adjusted (Supplementary Table S5; basic Model 1 for age, sex, and BMI; and extra Model 2 for age, sex, BMI, history of hypercholesterolemia, myocardial infarction, and statin therapy), neither whole blood nor blood mononuclear cell samples were significantly differentially expressed by CAD status. These results provide some evidence against meaningful changes in blood complement system mRNA levels during atherosclerosis.

### Cluster analysis of arterial, whole blood, and mononuclear samples

The cluster analysis results are presented in Figure 3 and Supplementary Table S7. UMAP over arteries indicates that plaques and normal arteries have clearly separate profiles. There is also a small subcluster in the plaques, but this has no self-evident connection to the AHA class. The UMAPs of the blood samples confirmed that the samples were similar in terms of complement transcripts. WGCNA assigns approximately half of the transcripts into a single weak module that can be weakly associated with the difference between plaques and normal arteries. More interestingly, the most connected, central genes in this TOM network include *FPR3, C1QC, ITGB2, ITGB2, ITGAM,* and *C1QB*. Overall, these results are consistent with a simple scenario in which mostly the differentially expressed transcripts form a single cluster that is expressed differently between all plaques and normal arteries.

**Figure 3.**
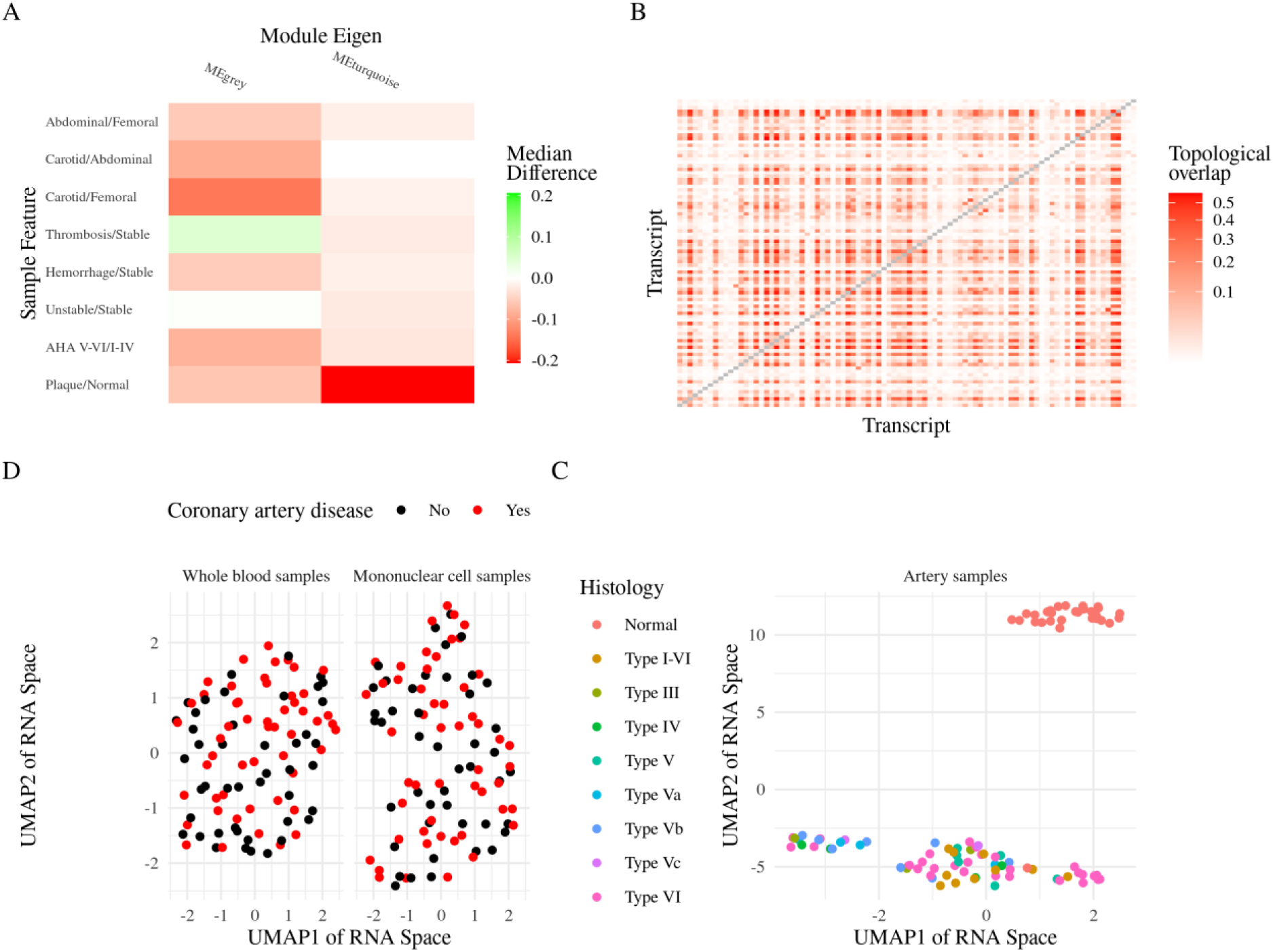
A. Heatmap showing associations of plaque features and WGCNA-predicted module eigenvectors. B. Adjacency matrix of the WGCNA network with hierarchical clustering of ordered rows and columns. C. UMAP clustering of the artery samples. D. UMAP clustering of whole blood and blood mononuclear cell samples.

### Gene Set Enrichment Analysis of the Gene Ontology and KEGG knowledge bases

Supplementary Table 8 presents the enriched Gene Ontology sets, and Supplementary Tables S9- 10 present all the results for the 19 KEGG pathways and enriched Gene Ontology sets per arterial bed, which are based on the set of all transcripts (90 genes). Additionally, Venn diagrams comparing the results from subgroup analyses by vessel are presented in Supplementary Figures S4-5.

In summary, 287 GO terms and 12 KEGG pathways were enriched with differentially expressed genes at the FDR<0.25 (mGSZ) level, and 41 GO terms and 9 KEGG pathways were enriched at the FDR<0.10 (mGSZ) level. A total of 7.4% of all the GO terms were enriched in all the arteries separately.

The Gene Ontology results suggest that the differential complement expression pattern observed in the plaques could be due to processes related to cell junctions and leukocyte, macrophage, lymphocyte, and T-cell activation, as well as phagocytosis, cell killing, apoptotic cell clearance, secretion, and protein localization. On the other hand, regulation of complement-dependent cytotoxicity (five genes) was enriched, with an mGSZ FDR of 0.13. In KEGG, complement and coagulation cascades are clearly enriched (mGSZ FDR 0.076), but this seems to be weakly shifted expression (odds ratio 1.1).

### Single-cell RNA-Seq location analysis of the top transcripts

The largest predicted cell type was macrophages, with 2,039 cells, and the remaining predicted cell types were fibroblasts (2 subtypes), smooth muscle cells (2 subtypes), endothelial cells (2 subtypes), T cells, B cells, plasma cells, Schwann cells, and mast cells. A reanalysis of the macrophages revealed four macrophage subclusters, but the expression of the top transcripts was similar in all of them (Supplemental Figure S6).

The expression of the top atherosclerosis-associated (AHA I-VI plaques vs. normal arteries) genes in single cells is presented in Figure 4. First, six genes were expressed in one cell type, namely- *C3AR1, C5AR1, FPR3, ITGAM*, and *TREM2* in macrophages and *C6* in fibroblasts. Two additional genes (*C1QB* and *C1QC*) are highly expressed, mostly in macrophages. On the other hand, two genes (*C5* and *GPLD1*) have very low expression and cannot be confidently localized. The expression of the ten remaining genes was more complicated. *C1S*, *MFAP4*, and *SERPING1* are expressed mostly in nonimmune cells. *CD46*, *CD55*, and PHB are expressed fairly uniformly across cell types. *ITGB2* is almost exclusive to macrophages and T cells, and *CD93,* encoding the C1q receptor subcomponent C1qRp, is expressed mostly in macrophages and endothelial cells. *C3* is highly expressed in fibroblasts but is expressed at low levels in Schwann cells and macrophages. Finally, *CFD* is widely expressed, with the highest levels in fibroblasts and macrophages. In terms of expression levels, the most relevant genes might include the gene encoding the C1 protein and C3 and the respective anaphylatoxin receptor, *CFD*, *ITGB2*, *SERPING1* and *MFAP4*.

**Figure 4.**
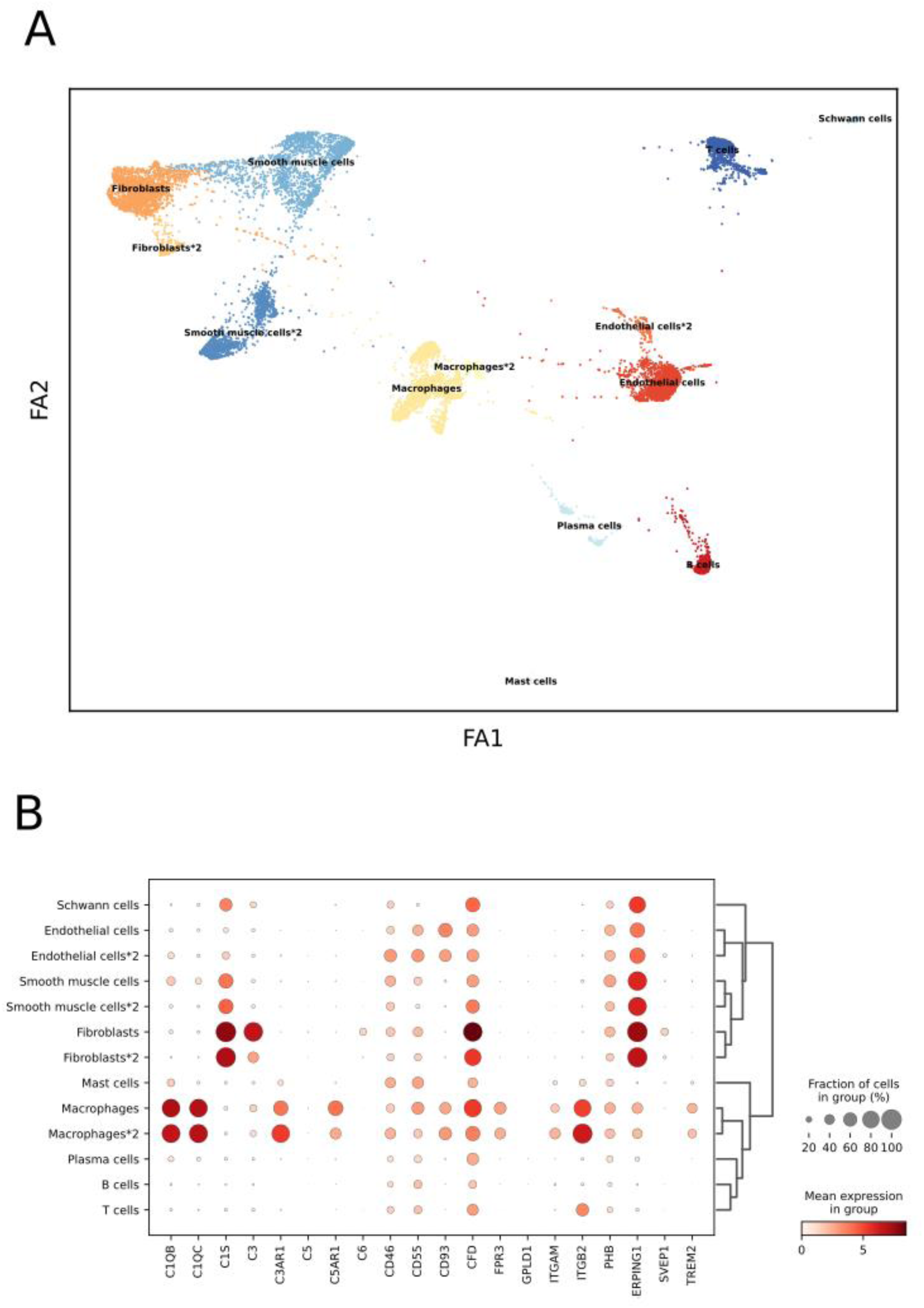
A. Clustering and annotation of single cells from plaque samples into major cell types. B. Expression of the top atherosclerosis-associated (AHA I-VI plaques vs. normal arteries) genes in single cells of each of the major cell types.

### Top gene correlations with different types of inflammatory and cell markers

The marker correlations for the top transcripts are shown in Figure 5, and those for all transcripts are shown in Supplementary Figure S7. We observed plaques that expressed higher levels of *TREM2*, CD93, *C5AR1, ITGAM, ITGB2, CiQB, C1QC, C3AR1, FPR3,* and *CFD* also tended to express higher levels of markers of M2 macrophages and inflammation. Conversely, plaques that expressed higher levels of *C5, SERPING1, GPLD1* and *MFAP4* tended to have higher levels of markers of smooth muscle cells and SMC-rich plaques. Other genes have less consistent profiles.

**Figure 5.**
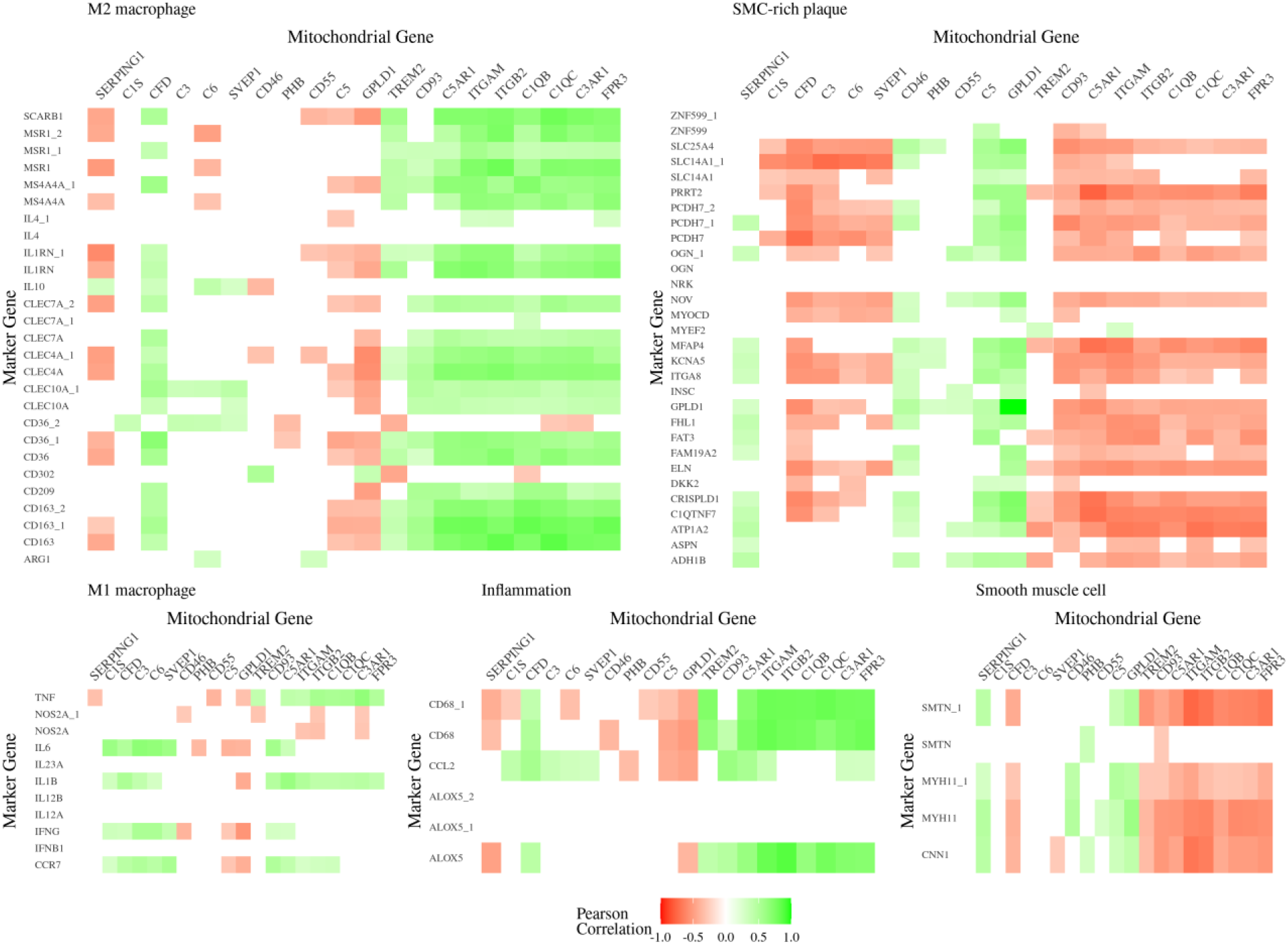
Heatmaps summarizing the Pearson correlation structure between the top RNA transcripts and cell marker RNA transcripts, named with their corresponding gene symbols. Green indicates a positive correlation, and red indicates a negative correlation. Only statistically significant (Benjamini‒Hochberg FDR<0.05) Pearson correlations are shown for clarity. SMC, smooth muscle cell.

These results are mostly consistent with single-cell localization; however, some differences exist. While *C5* and *GPLD1* were less clear in the single-cell results, this simple correlation analysis suggested that they were mostly localized to smooth muscle-rich areas. In fact, *GPLD1* is an SMC-rich marker. For *C1S*, the marker correlations do not support non-immune localization much, as the correlations are very low. Finally, marker correlations suggest that *CFD* is expressed more alongside inflammatory markers than smooth muscle markers.

## Discussion

In summary, over one-third of the complement system genes are differentially expressed in plaques and are mostly upregulated, but different stages of plaque and blood samples are markedly similar. Only a few genes presented clear vessel differences, but the gene set enrichment results were highly dependent on the analyzed vessel.

*CFD* was one of the most downregulated genes of the complement system in plaques but was still highly expressed in all plaque cell types, especially macrophages. *CFD* was also barely upregulated in thrombotic plaques compared with stable plaques. Factor D is a serine protease that enables the activation of the alternative pathway and thus affects opsonization, phagocytosis, and ultimately target cell lysis by the membrane attack complex^32^. Few reports exist on CFD (also known as adipsin) in atherosclerosis directly. Among the 50 cytokines, blood CFD (adipsin) was the best predictor of death in coronary artery disease patients^33^. Similarly, blood adipsin is a decent predictor of carotid intimal thickness in polycystic ovary syndrome patients^34^ and obese adults^35^. In ApoE-negative mice, transgenic adipsin reduced the plaque area and degree of macrophage deposition, and in macrophages in vitro, CFD overexpression decreased lipid accumulation in response to oxLDL^36^. On the other hand, knocking out adipsin in LDLR-negative mice did not affect lesion size or basic metabolic features^37^. The present results are consistent with the concept that all types of plaques locally under-express factor D in a majority of cell types.

*C6* was the most downregulated gene of the complement system in plaques, especially in carotid plaques, and was clearly localized to plaque fibroblasts. Previously, genetic C6 deficiency has been associated with resistance to diet-induced atherosclerosis in rabbits^38^, but in humans, only infectious and autoimmune cases related to C6 deficiency have been reported^39^. As the C6 protein is part of the membrane attack complex in the terminal complement pathway, the present results suggest diminished terminal pathway activation even in the presence of the upregulation of early complement pathways, suggesting an inflammatory mechanism in humans.

The present results suggest that *ITGB2* is an influential inflammatory gene in atherosclerosis. *ITGB2* (encoding CD18) is one of the most upregulated complement system genes in all plaques and is concentrated in macrophages and T cells, and together with β-integrin CD11b (encoded by *ITGAM*) or CD11c (encoded by *ITGAX*), it forms the complement receptor CR3 or CR4, respectively, whose main ligand is C3b/iC3b. This regulator of inflammation is one of the most central in the expression network. The cell junction-related gene sets were also among the most enriched. *ITGB2* was previously identified as one of the top atherosclerosis genes on the basis of an analysis of their protein interaction network^40^, and it was also identified as one of the key genes associated with intraplaque hemorrhage^41^. Integrins are well known to have a wide range of effects on atherosclerosis, especially on endothelial function, angiogenesis, leukocyte targeting and functions, and smooth muscle functions (migration, proliferation) in plaques, as well as platelet function during atherothrombosis^42^. In plaques rich in SMCs, the downregulation of alternative pathway activators *CFD* and *C3* and classical pathway activators *C1qB* & -*C*, *C1S* and complement receptors (*SERPING1* encoding C1inh, anaphylatoxin receptors *C3aR*, *C5aR* and *ITGAM* & *ITGB2* encoding CR3) concomitantly with the upregulation of complement inhibitors *CD46* and *CD55* indicates diminished complement activation in these plaques. MFAP4, encoding microfibrillar-associated protein 4, is also associated with SMC-rich plaques and is a known integrin-dependent promoter of vascular smooth muscle migration and monocyte chemotaxis^43^. Plaques characterized by inflammation and macrophage populations present an opposite pattern, with upregulation of *CFD* and complement receptors and downregulation of surface-bound complement inhibitors. The Gene Ontology association data corroborate these observations, where few canonical complement functions, including phagocytosis and cell death regulation, are implicated; however, cross effects on adaptive immune activation through associations with cell‒ cell signaling, cell activation and activation of the leukocyte response are highlighted. Together, these results indicate that complement has a local regulatory effect on atherosclerosis. These findings suggest that, depending on the histopathology of a particular plaque, complement may have diverse functions in driving the pathogenesis of atherosclerosis.

Major earlier studies have generally focused on only one arterial bed (most commonly carotid artery plaques); therefore, the Tampere Vascular Study was originally designed to cover four atherosclerotic vascular beds from different regions of the artery tree and compare these to samples from which atherosclerosis was not detected via histology. This allows comparative vascular region expression studies; however, Tampere Vascular Study is statistically powered mainly for hypothesis-driven research of plausible genes, and statistical power becomes low in regionwise plaque subgroup analyses. Owing to ethical limitations, we were not able to obtain any corresponding normal arteries from the carotid, aortic, or femoral regions as controls. Therefore, vascular samples of atherosclerotic lesions are from different arteries than control samples are, and the association between gene expression and the presence of atherosclerosis might be confounded by the type of artery. A more accurate analysis could be performed between healthy subjects and those with atherosclerosis. Because of the systemic nature of atherosclerosis, internal thoracic artery samples from patients subjected to coronary artery bypass grafting may be affected by atherosclerosis and the same risk factors as other vascular beds, although no microscopic or histological evidence of plaques in major LITAs could be demonstrated. Owing to vascular sampling, the measurement of histopathology and RNA expression is always cross-sectional, which also affects causal interpretations. From a methodological point of view, the reliability of the genome-wide expression (GWE) results was confirmed in our previous study by comparing the GWE results with those of the TaqMan quantitative LDA qRT‒PCR method^19^. In comparison, the Pearson correlation coefficient between methods was 0.87, indicating that the fold changes were in good agreement, although for highly up- or downregulated transcripts, GWE yielded lower absolute fold change values than did LDA.

Taken together, the present findings support the hypothesis that dysregulation of gene expression is associated with atherosclerosis development and that there are vascular regional differences in the local role of the complement system in atherosclerosis. The varying histomorphology of plaque tissue is characterized by inflammatory processes distinguishable by differential gene expression. Furthermore, our results support the local effect of complement-mediated inflammation in inflammatory plaque tissue but not in SMC-dominated plaques.

## Data Availability

The dataset supporting the conclusions of this article were obtained from the Tampere Vascular study which comprises health related participant data. The use of data is restricted under the regulations on professional secrecy (Act on the Openness of Government Activities, 612/1999) and on sensitive personal data (Personal Data Act, 523/1999, implemented in the EU data protection directive 95/46/EC). Due to these restrictions, the data cannot be stored in public repositories or otherwise made publicly available. Data access may be permitted on a case-by-case basis upon request only. Data sharing outside the group is done in collaboration with the TVS group and requires a data-sharing agreement. Investigators can submit an expression of interest to the chairman of the publication committee (Prof Terho Lehtimäki, Tampere University, Finland).

## Ethical approval

The Tampere Vascular Study (TVS) (ETL-code R99204) and Finnish Cardiovascular Study (FINCAVAS) (ETL-code R00153) were approved by the Ethics Committee of Tampere Hospital District. All studies were conducted in accordance with the Declaration of Helsinki, and all study subjects provided written informed consent.

## Data availability

The dataset supporting the conclusions of this article was obtained from the Tampere Vascular study, which comprises health-related participant data. The use of data is restricted under the regulations on professional secrecy (Act on the Openness of Government Activities, 612/1999) and sensitive personal data (Personal Data Act, 523/1999, implemented in the EU data protection directive 95/46/EC). Owing to these restrictions, the data cannot be stored in public repositories or otherwise made publicly available. Data access may be permitted on a case-by-case basis upon request only. Data sharing outside the group is performed in collaboration with the TVS group and requires a data-sharing agreement. Investigators can submit an expression of interest to the chairperson of the publication committee (Prof. Terho Lehtimäki, Tampere University, Finland).

## Acknowledgments and funding

This study was supported by grants from the Competitive Research Funding of the Tampere University Hospital (Grant 9M048, 9N035, X51001 for T.L.), Emil Aaltonen Foundation (T.L.), Pirkanmaa Regional Fund of the Finnish Cultural Foundation, the Research Foundation of Orion Corporation, the Jenny and Antti Wihuri Foundation, Research Council of Finland (grants 322098 (E.R.) and 104821), Finnish Foundation for Cardiovascular Research, Yrjö Jahnsson Foundation, European Union 7th Framework Program (grant 201668 for AtheroRemo), EU Horizon 2020 (grant 755320 for TAXINOMISIS and grant 848146 for To Aition), Tampere University Hospital Supporting Foundation, the Finnish Society of Clinical Chemistry, and the Wellbeing Services County of Pirkanmaa and Fimlab Laboratoriot Oy.

## Author contributions

E.T. performed the microarray and scRNA-Seq data analysis and wrote the first draft of the manuscript. A.I.L. participated in the data interpretation, finalized the manuscript and oversaw the editorial process. N.O. participated in the data collection and conceptualization and reviewed the manuscript. H.B. developed the scRNA-Seq data analysis and reviewed the manuscript. S.Ma. and E.R. acquired resources, performed sample analysis and reviewed the manuscript; L-P.L. and P.P.M. helped with the statistical analysis and reviewed the manuscript; I.K. performed the histopathology analysis and reviewed the manuscript; S.P. and A.M. acquired data and resources and reviewed the manuscript; N.M. participated in supervision and reviewed the manuscript; and M.K. handled funding, participated in cohort collection and reviewed the manuscript. S.Me. participated in conceptualization of the study and reviewed the manuscript; S.P. and T.L. handled funding, supervision, conceptualization, and reviewed the manuscript.

## References

1. Roth GA, Abate D, Abate KH, Abay SM, Abbafati C, Abbasi N, Abbastabar H, Abd-Allah F, Abdela J, Abdelalim A, Abdollahpour I, Abdulkader RS, Abebe HT, Abebe M, Abebe Z, et al. Global, regional, and national age-sex-specific mortality for 282 causes of death in 195 countries and territories, 1980–2017: a systematic analysis for the Global Burden of Disease Study 2017. The Lancet. 2018;392(10159):1736–1788.

2. Libby P, Buring JE, Badimon L, Hansson GK, Deanfield J, Bittencourt MS, Tokgözoğlu L, Lewis EF. Atherosclerosis. Nature Reviews Disease Primers. 2019;5(1).

3. Merle NS, Church SE, Fremeaux-Bacchi V, Roumenina LT. Complement system part I - molecular mechanisms of activation and regulation. Frontiers in Immunology. 2015;6(JUN).

4. Merle NS, Noe R, Halbwachs-Mecarelli L, Fremeaux-Bacchi V, Roumenina LT. Complement system part II: Role in immunity. Frontiers in Immunology. 2015;6(MAY).

5. Lokki AI, Heikkinen-Eloranta J. Pregnancy induced TMA in severe preeclampsia results from complement-mediated thromboinflammation. Human Immunology. 2021;82(5):371–378.

6. Vlaicu SI, Tatomir A, Rus V, Mekala AP, Mircea PA, Niculescu F, Rus H. The role of complement activation in atherogenesis: the first 40 years. Immunologic Research. 2016;64(1):1–13.

7. Oksjoki R, Jarva H, Kovanen PT, Laine P, Meri S, Pentikäinen MO. Association between complement factor H and proteoglycans in early human coronary atherosclerotic lesions: Implications for local regulation of complement activation. *Arteriosclerosis*, Thrombosis, and Vascular Biology. 2003;23(4):630–636.

8. Yasojima K, Schwab C, McGeer EG, McGeer PL. Complement components, but not complement inhibitors, are upregulated in atherosclerotic plaques. *Arteriosclerosis*, Thrombosis, and Vascular Biology. 2001;21(7):1214–1219.

9. Cao W, Bobryshev Y V., Lord RSA, Oakley REI, Lee SH, Lu J. Dendritic cells in the arterial wall express C1q: Potential significance in atherogenesis. Cardiovascular Research. 2003;60(1):175–186.

10. Niculescu F, Rus HG, Vlaicu R. Immunohistochemical localization of C5b-9, S-protein, C3d and apolipoprotein B in human arterial tissues with atherosclerosis. Atherosclerosis. 1987;65(1–2):1–11.

11. Martínez-López D, Roldan-Montero R, García-Marqués F, Nuñez E, Jorge I, Camafeita E, Minguez P, Rodriguez de Cordoba S, López-Melgar B, Lara-Pezzi E, Fernández-Ortiz A, Ibáñez B, Valdivielso JM, Fuster V, Michel JB, et al. Complement C5 Protein as a Marker of Subclinical Atherosclerosis. Journal of the American College of Cardiology. 2020;75(16):1926– 1941.

12. Kiss MG, Binder CJ. The multifaceted impact of complement on atherosclerosis. Atherosclerosis. 2022;351:29–40.

13. Oksala N, Pärssinen J, Seppälä I, Raitoharju E, Ivana K, Hernesniemi J, Lyytikäinen LP, Levula M, Mäkelä KM, Sioris T, Kähönen M, Laaksonen R, Hytönen V, Lehtimäki T. Association of neuroimmune guidance cue netrin-1 and its chemorepulsive receptor UNC5B with atherosclerotic plaque expression signatures and stability in human(s) Tampere Vascular Study (TVS). Circulation: Cardiovascular Genetics. 2013;6(6):579–587.

14. Sulkava M, Raitoharju E, Levula M, Seppälä I, Lyytikaïnen LP, Mennander A, Järvinen O, Zeitlin R, Salenius JP, Illig T, Klopp N, Mononen N, Laaksonen R, Kähönen M, Oksala N, et al. Differentially expressed genes and canonical pathway expression in human atherosclerotic plaques-Tampere Vascular Study. Scientific Reports. 2017;7.

15. Nieminen T, Lehtinen R, Viik J, Lehtimäki T, Niemelä K, Nikus K, Niemi M, Kallio J, Kööbi T, Turjanmaa V, Kähönen M. The Finnish Cardiovascular Study (FINCAVAS): Characterising patients with high risk of cardiovascular morbidity and mortality. BMC Cardiovascular Disorders. 2006;6.

16. Stary HC, Blankenhorn DH, Bleakley Chandler A, Glagov S, Insull W, Richardson M, Rosenfeld ME, Schaffer SA, Schwartz CJ, Wagner WD, Wissler RW. A definition of the intima of human arteries and of its atherosclerosis-prone regions: A report from the committee on vascular lesions of the council on arteriosclerosis, American Heart Association. *Arteriosclerosis*, Thrombosis, and Vascular Biology. 1992;12(1):120–134.

17. Anon. R Core Team (2021) R A Language and Environment for Statistical Computing. R Foundation for Statistical Computing, Vienna. - References - Scientific Research Publishing.

18. Du P, Kibbe WA, Lin SM. lumi: A pipeline for processing Illumina microarray. Bioinformatics. 2008;24(13):1547–1548.

19. Raitoharju E, Seppälä I, Lyytikäinen LP, Levula M, Oksala N, Klopp N, Illig T, Laaksonen R, Kähönen M, Lehtimäki T. A comparison of the accuracy of Illumina HumanHT-12 v3 Expression BeadChip and TaqMan qRT-PCR gene expression results in patient samples from the Tampere Vascular Study. Atherosclerosis. 2013;226(1):149–152.

20. Ashburner M, Ball CA, Blake JA, Botstein D, Butler H, Cherry JM, Davis AP, Dolinski K, Dwight SS, Eppig JT, Harris MA, Hill DP, Issel-Tarver L, Kasarskis A, Lewis S, et al. Gene ontology: Tool for the unification of biology. Nature Genetics. 2000;25(1):25–29.

21. Mann HBWDR. On a Test of Whether one of Two Random Variables is Stochastically Larger than the Other. The Annals of Mathematical Statistics. 1947;18(1):50–60.

22. Benjamini Y, Hochberg Y. Controlling the False Discovery Rate: A Practical and Powerful Approach to Multiple Testing. Journal of the Royal Statistical Society. Series B (Methodological). 1995;57(1):289–300.

23. Puig O, Yuan J, Stepaniants S, Zieba R, Zycband E, Morris M, Coulter S, Yu X, Menke J, Woods J, Chen F, Ramey DR, He X, O’Neill EA, Hailman E, et al. A gene expression signature that classifies human atherosclerotic plaque by relative inflammation status. Circulation: Cardiovascular Genetics. 2011;4(6):595–604.

24. Subramanian A, Tamayo P, Mootha VK, Mukherjee S, Ebert BL, Gillette MA, Paulovich A, Pomeroy SL, Golub TR, Lander ES, Mesirov JP. Gene set enrichment analysis: A knowledge-based approach for interpreting genome-wide expression profiles. Proceedings of the National Academy of Sciences of the United States of America. 2005;102(43):15545–15550.

25. Wolf FA, Angerer P, Theis FJ. SCANPY: Large-scale single-cell gene expression data analysis. Genome Biology. 2018;19(1).

26. Clough E, Barrett T. The Gene Expression Omnibus database. Methods in Molecular Biology. 2016;1418:93–110.

27. Wirka RC, Wagh D, Paik DT, Pjanic M, Nguyen T, Miller CL, Kundu R, Nagao M, Coller J, Koyano TK, Fong R, Woo YJ, Liu B, Montgomery SB, Wu JC, et al. Atheroprotective roles of smooth muscle cell phenotypic modulation and the TCF21 disease gene as revealed by single-cell analysis. Nature Medicine. 2019;25(8):1280–1289.

28. Zheng GXY, Terry JM, Belgrader P, Ryvkin P, Bent ZW, Wilson R, Ziraldo SB, Wheeler TD, McDermott GP, Zhu J, Gregory MT, Shuga J, Montesclaros L, Underwood JG, Masquelier DA, et al. Massively parallel digital transcriptional profiling of single cells. Nature Communications. 2017;8.

29. Cao Y, Wang X, Peng G. SCSA: A cell type annotation tool for single-cell RNA-seq data. Frontiers in Genetics. 2020;11.

30. Zhang X, Lan Y, Xu J, Quan F, Zhao E, Deng C, Luo T, Xu L, Liao G, Yan M, Ping Y, Li F, Shi A, Bai J, Zhao T, et al. CellMarker: A manually curated resource of cell markers in human and mouse. Nucleic Acids Research. 2019;47(D1):D721–D728.

31. Franzén O, Gan LM, Björkegren JLM. PanglaoDB: A web server for exploration of mouse and human single-cell RNA sequencing data. Database. 2019;2019(1).

32. Barratt J, Weitz I. Complement Factor D as a Strategic Target for Regulating the Alternative Complement Pathway. Frontiers in Immunology. 2021;12.

33. Ohtsuki T, Satoh K, Shimizu T, Ikeda S, Kikuchi N, Satoh T, Kurosawa R, Nogi M, Sunamura S, Yaoita N, Omura J, Aoki T, Tatebe S, Sugimura K, Takahashi J, et al. Identification of Adipsin as a Novel Prognostic Biomarker in Patients With Coronary Artery Disease. Journal of the American Heart Association. 2019;8(23).

34. Gursoy Calan O, Calan M, Yesil Senses P, Unal Kocabas G, Ozden E, Sari KR, Kocar M, Imamoglu C, Senses YM, Bozkaya G, Bilgir O. Increased adipsin is associated with carotid intima media thickness and metabolic disturbances in polycystic ovary syndrome. Clinical Endocrinology. 2016;85(6):910–917.

35. Zhang J, Teng F, Pan L, Guo D, Liu J, Li K, Yuan Y, Li W, Zhang H. Circulating adipsin is associated with asymptomatic carotid atherosclerosis in obese adults. BMC Cardiovascular Disorders. 2021;21(1).

36. Duan Y, Zhang X, Zhang X, Lin J, Shu X, Man W, Jiang M, Zhang Y, Wu D, Zhao Z, Sun D. Inhibition of macrophage-derived foam cells by Adipsin attenuates progression of atherosclerosis. Biochimica et Biophysica Acta - Molecular Basis of Disease. 2022;1868(12).

37. Liu L, Chan M, Yu L, Wang W, Qiang L. Adipsin deficiency does not impact atherosclerosis development in Ldlr―/― mice. American Journal of Physiology - Endocrinology and Metabolism. 2021;320(1):E87–E92.

38. Schmiedt W, Kinscherf R, Deigner HP, Kamencic H, Nauen O, Kilo J, Oelert H, Metz J, Bhakdi S. Complement C6 deficiency protects against diet-induced atherosclerosis in rabbits. *Arteriosclerosis*, Thrombosis, and Vascular Biology. 1998;18(11):1790–1795.

39. Pettigrew HD, Teuber SS, Gershwin ME. Clinical significance of complement deficiencies. Annals of the New York Academy of Sciences. 2009;1173:108–123.

40. Banik SK, Baishya S, Das Talukdar A, Choudhury MD. Network analysis of atherosclerotic genes elucidates druggable targets. BMC Medical Genomics. 2022;15(1).

41. Li S, Zhang Q, Huang Z, Tao W, Zeng C, Yan L, Chen F. Comprehensive analysis of immunocyte infiltration and the key genes associated with intraplaque hemorrhage in carotid atherosclerotic plaques. International Immunopharmacology. 2022;106.

42. Finney AC, Stokes KY, Pattillo CB, Orr AW. Integrin signaling in atherosclerosis. Cellular and Molecular Life Sciences. 2017;74(12):2263–2282.

43. Schlosser A, Pilecki B, Hemstra LE, Kejling K, Kristmannsdottir GB, Wulf-Johansson H, Moeller JB, Füchtbauer EM, Nielsen O, Kirketerp-Møller K, Dubey LK, Hansen PBL, Stubbe J, Wrede C, Hegermann J, et al. MFAP4 Promotes Vascular Smooth Muscle Migration, Proliferation and Accelerates Neointima Formation. *Arteriosclerosis*, Thrombosis, and Vascular Biology. 2016;36(1):122–133.

